# Comparative Analysis of Short-Chain Fatty Acid Levels as Predictors of Intestinal Mucosal Resilience in Pediatric Epilepsy Patients According to Duration of Valproic Acid Therapy

**DOI:** 10.1101/2025.07.31.25332438

**Authors:** Jeanette I. Ch. Manoppo, Praevilia M. Salendu, Hervina Yulanda

**Affiliations:** Department of Pediatrics, Faculty of Medicine, Sam Ratulangi University, Manado, Indonesia

**Keywords:** Short-chain fatty acids, valproic acid, pediatric epilepsy, intestinal mucosal resilience

## Abstract

**Background:** Epilepsy is the most common neurological disorder in children and often requires long-term treatment with valproic acid, which may affect intestinal mucosal resilience. Short-chain fatty acid (SCFA) levels can serve as predictors of gut mucosal integrity in pediatric epilepsy patients undergoing valproic acid therapy.

**Methods:** This cross-sectional study was conducted at the Pediatric Outpatient Clinic, Department of Child Health, Prof. Dr. R.D. Kandou General Hospital, and Prodia Clinical Laboratory in Manado, from December 2023 to April 2024. Subjects were children with epilepsy aged 6 months to 18 years who met the inclusion criteria. SCFA levels—including total SCFA, acetic acid, propionic acid, and butyric acid—were measured using gas chromatography.

**Results:** Of the 56 participants, 28 children had received valproic acid therapy for less than one year, and 28 for more than one year. The <1-year therapy group showed significantly higher levels of total SCFA (MD = 4.67 [3.22–6.13]; p < 0.001), acetic acid (MD = 15.53 [12.65–18.41]; p < 0.001), propionic acid (MD = 6.25 [4.16–8.33]; p < 0.001), and butyric acid (MD = 9.53 [7.04–12.02]; p < 0.001) compared to the >1-year therapy group. A statistically significant difference in SCFA levels was observed between the two groups (p < 0.05).

**Conclusion:** Duration of valproic acid therapy significantly affects SCFA levels in children with epilepsy.

## INTRODUCTION

Epilepsy is a common chronic neurological disorder in children, characterized by a predisposition to recurrent epileptic seizures. It affects more than 50 million individuals worldwide, including 0.5–1% of children [1]. The incidence is higher in developing countries due to suboptimal healthcare services in managing risk factors such as perinatal complications and infections [2]. Globally, the incidence of epilepsy is estimated at 102 per 100,000 per year, with the highest rate occurring during the first year of life and decreasing with age [3,4]. In Indonesia, epidemiological data are limited, as many cases remain undiagnosed [2]. Approximately 40% of pediatric epilepsy patients experience status epilepticus before the age of two, with 75% presenting with seizures as the initial symptom [2].

Diagnosing epilepsy is often challenging due to various seizure mimics. It is confirmed when a patient experiences ≥2 unprovoked seizures ≥24 hours apart or has clinical features consistent with an epilepsy syndrome [2]. Diagnostic support includes neuroimaging, EEG, and genetic testing, particularly in early-onset cases [5]. About one-third of epilepsy cases are attributable to brain injury, while the remainder are thought to be genetically related [6]. Early detection is essential to prevent recurrence, improve prognosis, and maintain quality of life [7,8].

There is a strong association between neurological disorders and gut microbiota (GM) dysbiosis, primarily mediated by the microbiota–gut–brain axis (MGBA) [9–13]. GM imbalance can influence neurotransmission, neuroinflammation, blood–brain barrier (BBB) permeability, and microglial function [14,15]. The human microbiota is established prenatally and distributed throughout various tissues, with the colon harboring the majority of microbial cells, estimated at 10^3^–10□ per gram [16,17].

Microbial metabolic products, such as short-chain fatty acids (SCFAs)—including acetate, propionate, and butyrate—play critical roles in maintaining intestinal epithelial integrity, immune regulation, and neurochemical modulation [18,19]. SCFAs are produced via fermentation of dietary fibers by colonic bacteria, with acetate being the predominant component [20]. The production of propionate and butyrate depends on specific metabolic pathways of certain bacterial groups, notably Bacteroidetes and Firmicutes [21].

Several studies have shown differences in gut microbial profiles between epilepsy patients and healthy controls [9–12]. For example, Xie et al. reported a predominance of Bacteroides and Bifidobacterium in healthy controls, while Firmicutes and Cronobacter were more abundant in epilepsy patients [22]. Huang et al. identified elevated levels of Streptococcus and Akkermansia in children with cerebral palsy and epilepsy (CPE) [23]. Gong et al. also demonstrated lower microbial diversity in epilepsy patients [24].

Valproic acid, a broad-spectrum antiepileptic drug (AED), has been a mainstay in epilepsy treatment for over 50 years [25]. Gut microbiota can influence drug biotransformation by modulating the expression of metabolic genes [26]. Valproic acid disrupts mitochondrial metabolism, induces oxidative stress, and alters gut microbial composition—elevating Actinobacteria and Firmicutes, while reducing Bacteroides and SCFA levels such as propionate and butyrate [27,28]. However, no studies in Indonesia have explored the relationship between valproic acid therapy duration and SCFA levels as markers of intestinal mucosal resilience in pediatric epilepsy.

Based on the aforementioned background, this study aims to investigate whether there is a difference in SCFA levels—used as predictors of intestinal mucosal resilience—in children with epilepsy based on the duration of valproic acid therapy. The null hypothesis (H□) states that there is no difference in SCFA levels based on therapy duration, whereas the alternative hypothesis (H□) posits that SCFA levels differ between patients treated with valproic acid for less than one year versus more than one year.

The general objective of this study is to evaluate and compare SCFA levels as indicators of intestinal mucosal resilience in pediatric epilepsy patients receiving valproic acid therapy. The specific objectives are: (1) to determine SCFA levels in children receiving valproic acid for less than one year, (2) to determine SCFA levels in children treated for more than one year, and (3) to compare SCFA levels between these two groups. This study is expected to provide several key contributions: (1) a clinical profile overview of pediatric epilepsy patients at Prof. Dr. R.D. Kandou General Hospital, Manado; (2) improved understanding of gut microbiota dysbiosis in pediatric epilepsy; and (3) scientific insights into SCFA levels as potential biomarkers in children receiving valproic acid therapy.

## METHODS

This study employed a cross-sectional design to assess differences in short-chain fatty acid (SCFA) levels among pediatric epilepsy patients treated with valproic acid for less than one year compared to those treated for more than one year. The study was conducted between December 2023 and April 2024 at the Pediatric Outpatient Clinic of Prof. Dr. R.D. Kandou General Hospital, Manado, and the Prodia Clinical Laboratory, Manado.

The study population consisted of children aged 6 months to 18 years diagnosed with epilepsy and receiving outpatient or inpatient care at the hospital. Participants were selected based on inclusion and exclusion criteria. Inclusion criteria were: (1) diagnosis of epilepsy, (2) receiving valproic acid monotherapy, and (3) informed consent obtained from parents. Exclusion criteria included ketogenic diet, recent use of probiotics/synbiotics, comorbid systemic diseases, or prior antibiotic use.

Sample size was determined using the following formula:

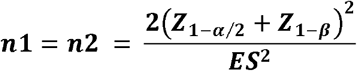

Notes:

n = required sample size

a = level of significance

z = quantile of the standard normal distribution

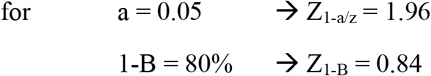

*effect size* (ES) = 80%

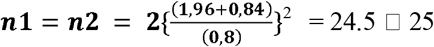

The calculated value of n1 was 25, resulting in a minimum required sample size of 50 participants for the study.

Ethical approval was obtained from the Ethics Committee of Prof. Dr. R.D. Kandou General Hospital. The independent variables in this study were SCFA levels (acetate, propionate, and butyrate), and the dependent variable was the duration of valproic acid therapy (<1 year vs. >1 year). Confounding variables controlled for included age, sex, seizure type and frequency, number of antiepileptic drugs, and comorbidities.

Fecal samples were collected using sterile containers and stored in a cool box at 2–8°C, with stability maintained for up to seven days. SCFA analysis was performed using gas chromatography-mass spectrometry (GC-MS). Sample preparation included homogenization, centrifugation, filtration, and the addition of ethyl butyrate as an internal standard.

Data analysis included descriptive statistics (mean, standard deviation, and frequency), bivariate analysis (independent t-test, one-way ANOVA, and Bonferroni post hoc test), and multivariate analysis using linear regression for each SCFA component. Statistical analysis was conducted using SPSS version 25.0 for Windows.

## RESULTS

### Respondent Characteristics

A total of 56 children with epilepsy were included in this study, consisting of 28 children who received valproic acid therapy for ≤1 year and 28 children who received therapy for >1 year.

Of the 56 participants, 30 were male (53.6%) and 26 were female (46.4%). The mean age of all respondents was 98.86 months, with children aged 1–3 years having a mean age of 22.45 months and those older than 3 years having a mean age of 115.04 months. The majority experienced generalized seizures (71.4%), and 71.4% had seizure durations of ≤2 minutes, with a mean duration of 1.33 minutes. For those with seizures lasting >2 minutes (28.6%), the mean duration was 6.81 minutes.

Regarding nutritional status, 75% had good nutritional status, 8.9% were undernourished, 10.7% were overweight, and 5.4% were obese. In terms of treatment duration with valproic acid, 17.9% had received therapy for 0–<3 months, and 12.5% had received therapy for >36 months.

Table 2 compares respondent characteristics between those receiving valproic acid therapy for ≤1 year and >1 year. Seizure types were evenly distributed in both groups (69.2% generalized, 30.8% focal). In terms of seizure duration, 64.3% of the ≤1 year group experienced seizures ≤2 minutes, compared to 78.6% in the >1 year group. Nutritional status showed that 67.9% of the ≤1 year group had good nutrition compared to 82.1% in the >1 year group. There were no statistically significant differences in age, seizure type, seizure duration, or nutritional status between the two groups.

**Table 1.** Respondent Characteristics in Children with Epilepsy.

| Variable | Total (n=56) |
| --- | --- |
| Sex, n (%) |  |
| Male | 30 (53.6) |
| Female | 26 (46.4) |
| Age (month), mean (sb) |  |
| Age (all participants) | 98.86 (55.76) |
| Age 1- 3 years old | 22.45 (5.82) |
| Age $> 3$ years old | 115.04 (46.45) |
| Seizure Type, n (%) |  |
| General | 40 (71.4) |
| Focal | 16 (28.6) |
| Seizure Duration, n (%) |  |
| $\leq 2$ minutes | 40 (71.4) |
| $> 2$ minutes | 16 (28.6) |
| Seizure Duration, mean (sb) |  |
| $\leq 2$ minutes | 1.33 (0.47) |
| $> 2$ minutes | 6.81 (6.50) |
| Nutritional Status, n (%) |  |
| Normal Nutrition | 42 (75.0) |
| Undernutrition | 5 (8.9) |
| Overweight | 6 (10.7) |
| Obesity | 3 (5.4) |
| Duration of valproic acid therapy, n(%) |  |
| 0 - 2 months | 10 (17.9) |
| 3 - 6 months | 5 (8.9) |
| 6 - 9 months | 4 (7.1) |
| 9 - 12 months | 9 (16.1) |
| 12 - 15 months | 2 (3.6) |
| 15 - 18 months | 2 (3.6) |
| 18 - 21 months | 2 (3.6) |
| 21 - 24 months | 4 (7.1) |
| 24 - 27 months | 3 (5.4) |
| 27 - 30 months | 2 (3.6) |
| 30 - 33 months | 2 (3.6) |
| 33 - 36 months | 4 (7.1) |
| $>36$ months | 7 (12.5) |

**Table 2.** Characteristics of Respondents by Duration of Valproic Acid Therapy.

| Variable | Valproic Acid Therapy |  |
| --- | --- | --- |
|  | ≤1 Year<br>(n=28) | > 1 Years<br>(n=28) |
| Seizure Type, n (%) |  |  |
| General | 18 (69.2) | 18 (69.2) |
| Focal | 8 (30.8) | 8 (30.8) |
| Seizure Duration, n (%) |  |  |
| ≤ 2 minutes | 18 (64.3) | 22 (78.6) |
| > 2 minutes | 10 (34.6) | 6 (21.4) |
| Nutritional Status |  |  |
| Normal Nutrition | 19 (67.9) | 23 (82.1) |
| Undernutrition | 4 (14.3) | 1 (3.6) |
| Overweight | 2 (7.1) | 4 (14.3) |
| Obesity | 3 (10.7) | 0 (0) |

### SCFA Profile by Duration of Valproic Acid Therapy

Table 3 and Figure 1 presents the mean SCFA levels based on therapy duration. Total SCFA (mg/dL) was significantly higher in the ≤1 year group (13.57 ± 2.82) compared to the >1 year group (8.89 ± 2.60), with a mean difference of 4.67 (95% CI: 3.22–6.13; p<0.001). Acetate, propionate, and butyrate percentages were also higher in the ≤1 year group (66.60%, 21.14%, 25.17%, respectively) than in the >1 year group (51.07%, 14.89%, 15.64%). Absolute butyrate (mg/dL) was also significantly higher in the ≤1 year group (3.13 vs. 1.79; mean difference 1.34, 95% CI: 0.99–1.68; p<0.001). These differences were statistically significant (p<0.001).

**Table 3.** Mean (SD) of SCFA Profiles by Valproic Acid Therapy Duration.

| Variable | Terapi Asam Valproat |  | Mean<br>Difference<br>(95% IK) | p<br>Value |
| --- | --- | --- | --- | --- |
|  | ≤1 Tahun<br>(n=28) | > 1 Tahun<br>(n=28) |  |  |
| Total SCFA<br>(mg/dL) | 13.57 (2.82) | 8.89 (2.60) | 4.67 (3.22:6.13) | <0.001 |
| Asam Asetat (%) | 66.60 (4.83) | 51.07<br>(5.89) | 15.53<br>(12.65:18.41) | <0.001 |
| Asam Propionat<br>(%) | 21.14 (3.81) | 14.89<br>(3.97) | 6.25 (4.16:8.33) | <0.001 |
| Asam Butirat (%) | 25.17 (4.84) | 15.64<br>(4.43) | 9.53 (7.04:12.02) | <0.001 |
| Asam Butirat<br>Absolut (mg/dL) | 3.13 (0.66) | 1.79 (0.62) | 1.34 (0.99:1.68) | <0.001 |

**Table 4.**
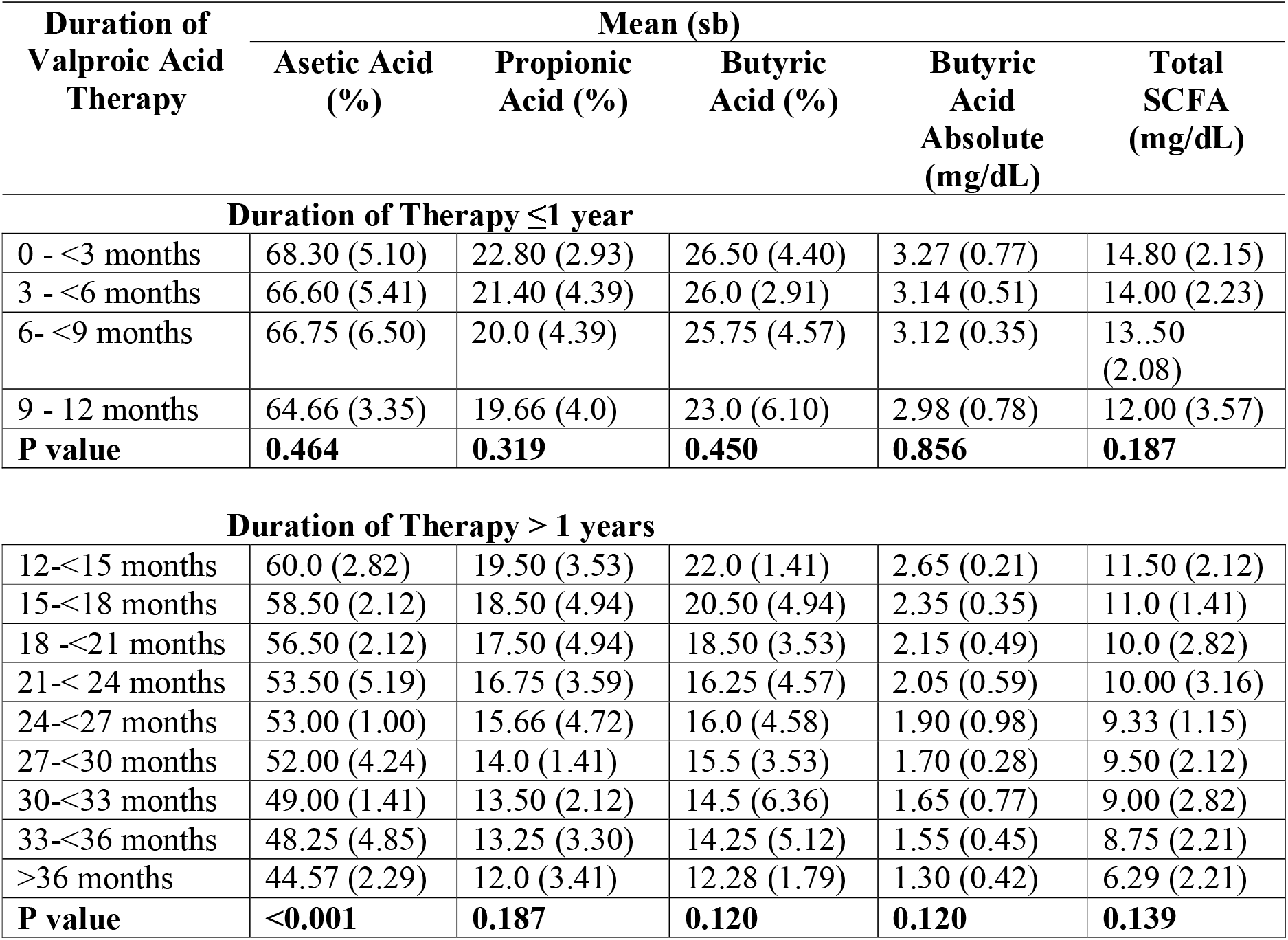
Within-Group SCFA Profile by Therapy Duration.

| Duration of<br>Valproic Acid<br>Therapy | Mean (sb) |  |  |  |  |
| --- | --- | --- | --- | --- | --- |
|  | Asetic Acid<br>(%) | Propionic<br>Acid (%) | Butyric<br>Acid (%) | Butyric<br>Acid<br>Absolute<br>(mg/dL) | Total<br>SCFA<br>(mg/dL) |
| **Duration of Therapy ≤1 year** |  |  |  |  |  |
| 0 - <3 months | 68.30 (5.10) | 22.80 (2.93) | 26.50 (4.40) | 3.27 (0.77) | 14.80 (2.15) |
| 3 - <6 months | 66.60 (5.41) | 21.40 (4.39) | 26.0 (2.91) | 3.14 (0.51) | 14.00 (2.23) |
| 6- <9 months | 66.75 (6.50) | 20.0 (4.39) | 25.75 (4.57) | 3.12 (0.35) | 13..50<br>(2.08) |
| 9 - 12 months | 64.66 (3.35) | 19.66 (4.0) | 23.0 (6.10) | 2.98 (0.78) | 12.00 (3.57) |
| **P value** | **0.464** | **0.319** | **0.450** | **0.856** | **0.187** |
| **Duration of Therapy > 1 years** |  |  |  |  |  |
| 12-<15 months | 60.0 (2.82) | 19.50 (3.53) | 22.0 (1.41) | 2.65 (0.21) | 11.50 (2.12) |
| 15-<18 months | 58.50 (2.12) | 18.50 (4.94) | 20.50 (4.94) | 2.35 (0.35) | 11.0 (1.41) |
| 18 -<21 months | 56.50 (2.12) | 17.50 (4.94) | 18.50 (3.53) | 2.15 (0.49) | 10.0 (2.82) |
| 21-< 24 months | 53.50 (5.19) | 16.75 (3.59) | 16.25 (4.57) | 2.05 (0.59) | 10.00 (3.16) |
| 24-<27 months | 53.00 (1.00) | 15.66 (4.72) | 16.0 (4.58) | 1.90 (0.98) | 9.33 (1.15) |
| 27-<30 months | 52.00 (4.24) | 14.0 (1.41) | 15.5 (3.53) | 1.70 (0.28) | 9.50 (2.12) |
| 30-<33 months | 49.00 (1.41) | 13.50 (2.12) | 14.5 (6.36) | 1.65 (0.77) | 9.00 (2.82) |
| 33-<36 months | 48.25 (4.85) | 13.25 (3.30) | 14.25 (5.12) | 1.55 (0.45) | 8.75 (2.21) |
| >36 months | 44.57 (2.29) | 12.0 (3.41) | 12.28 (1.79) | 1.30 (0.42) | 6.29 (2.21) |
| **P value** | **<0.001** | **0.187** | **0.120** | **0.120** | **0.139** |

**Table 5.**
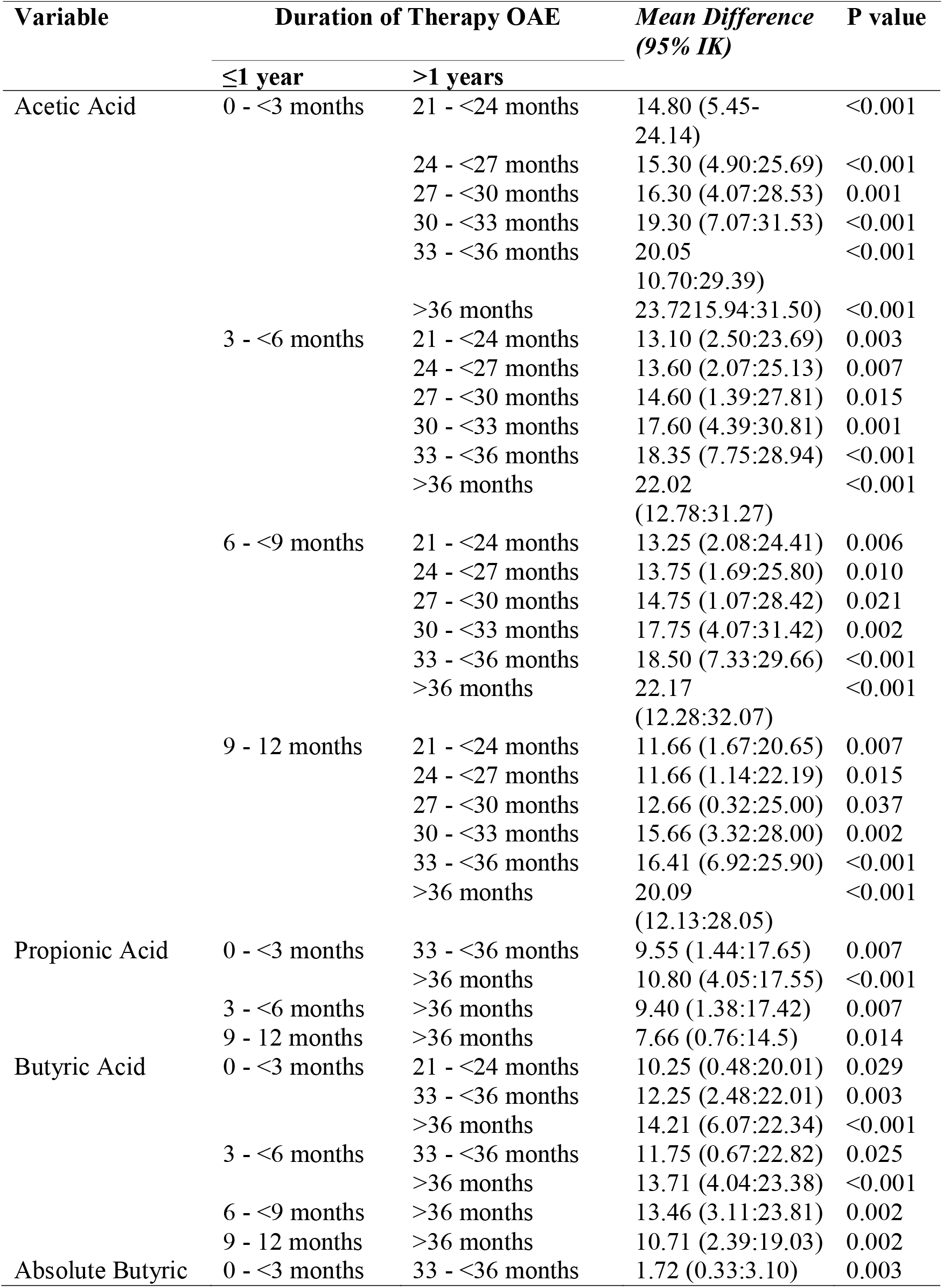
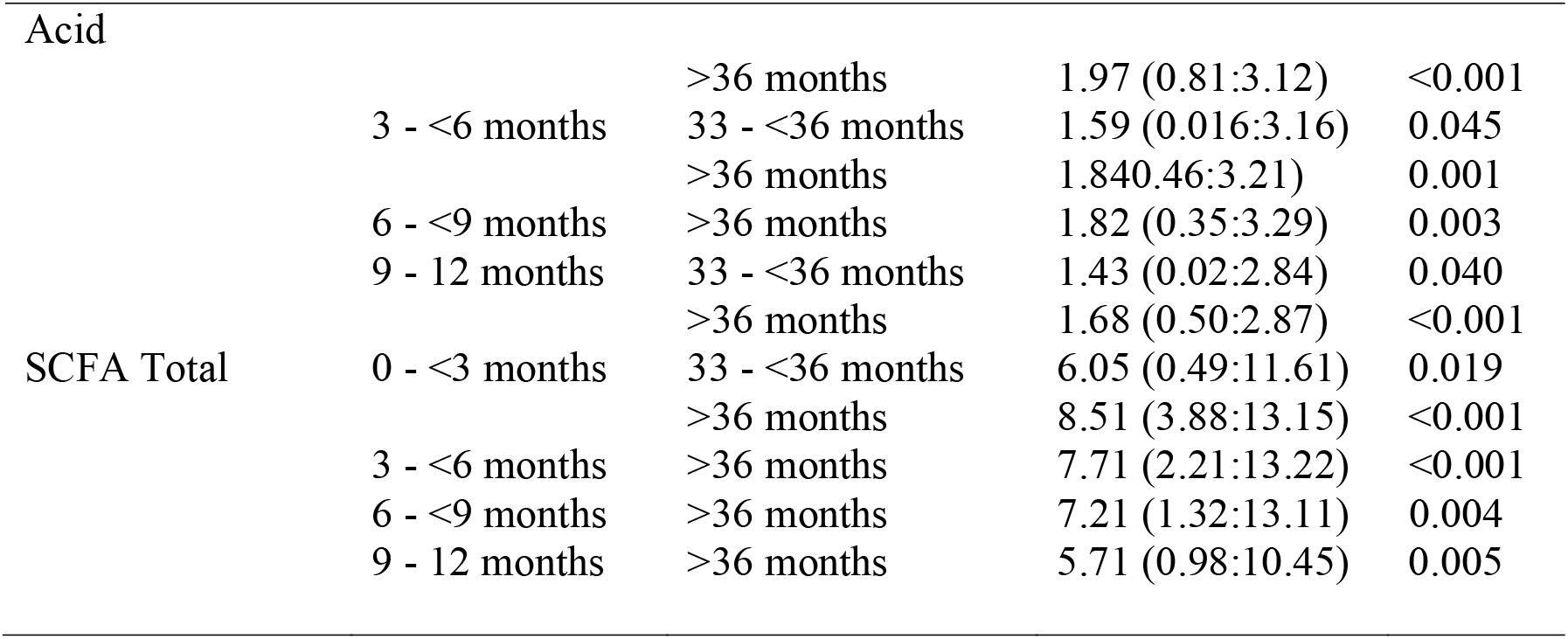
Statistically Significant SCFA Differences Between Duration Groups.

**Table 6.** Multivariate Analysis of Total SCFA and Duration of Therapy.

| Variable | OR (95% Confidence Interval) | p Value |
| --- | --- | --- |
| SCFA Total | 0.49 (0.33:0.73) | <0.001 |
| Age | 1.01 (0.98:1.01) | 0.832 |
| Seizure Type |  |  |
| General | 7.21 (1.03:50.25) | 0.046 |
| Focal | Reference |  |
| Seizure Duration |  |  |
| ≤2 minutes | Reference | 0.161 |
| >2 minutes | 3.41 (0.61:18.99) |  |

**Table 7.** Linear Regression Analysis of Short-Chain Fatty Acid Levels and Duration of Antiepileptic Therapy.

| Outcome | Univariate |  | Multivariate |  |
| --- | --- | --- | --- | --- |
| | $\beta$ (95% CI) | *p* | $\beta$ (95% CI) | *p* |
| Acetic Acid | -0.59 (-0.66 ; -0.51) | <0.001 | -0.58 (-0.66 ; -0.50) | <0.001 |
| Propionic Acid | -0.25 (-0.32 ; -0.19) | <0.001 | -0.26 (-0.32 ; -0.19) | <0.001 |
| Butyric Acid | -0.36 (-0.44 ; -0.28) | <0.001 | -0.37 (-0.46 ; -0.29) | <0.001 |
| Butyric Acid (absolute) | -0.05 (-0.06 ; -0.04) | <0.001 | -0.05 (-0.06 ; -0.03) | <0.001 |
| SCFA Total | -0.19 (-0.23 ; -0.14) | <0.001 | -0.18 (-0.22 ; -0.13) | <0.001 |

Note: CI = Confidence Interval. □ Multivariate model adjusted for valproic acid dosage variation, body weight, and their interaction.

**Figure 1.**
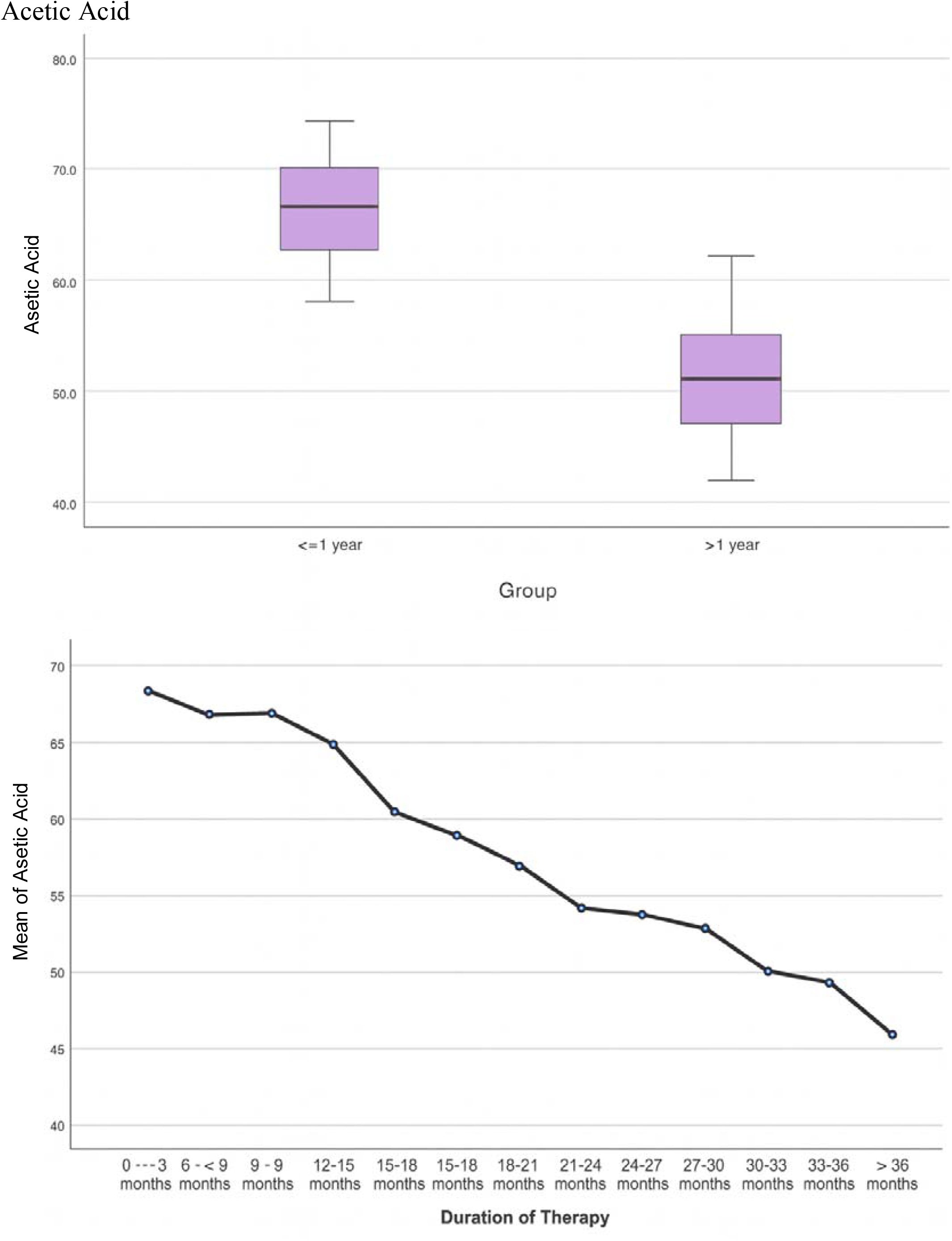
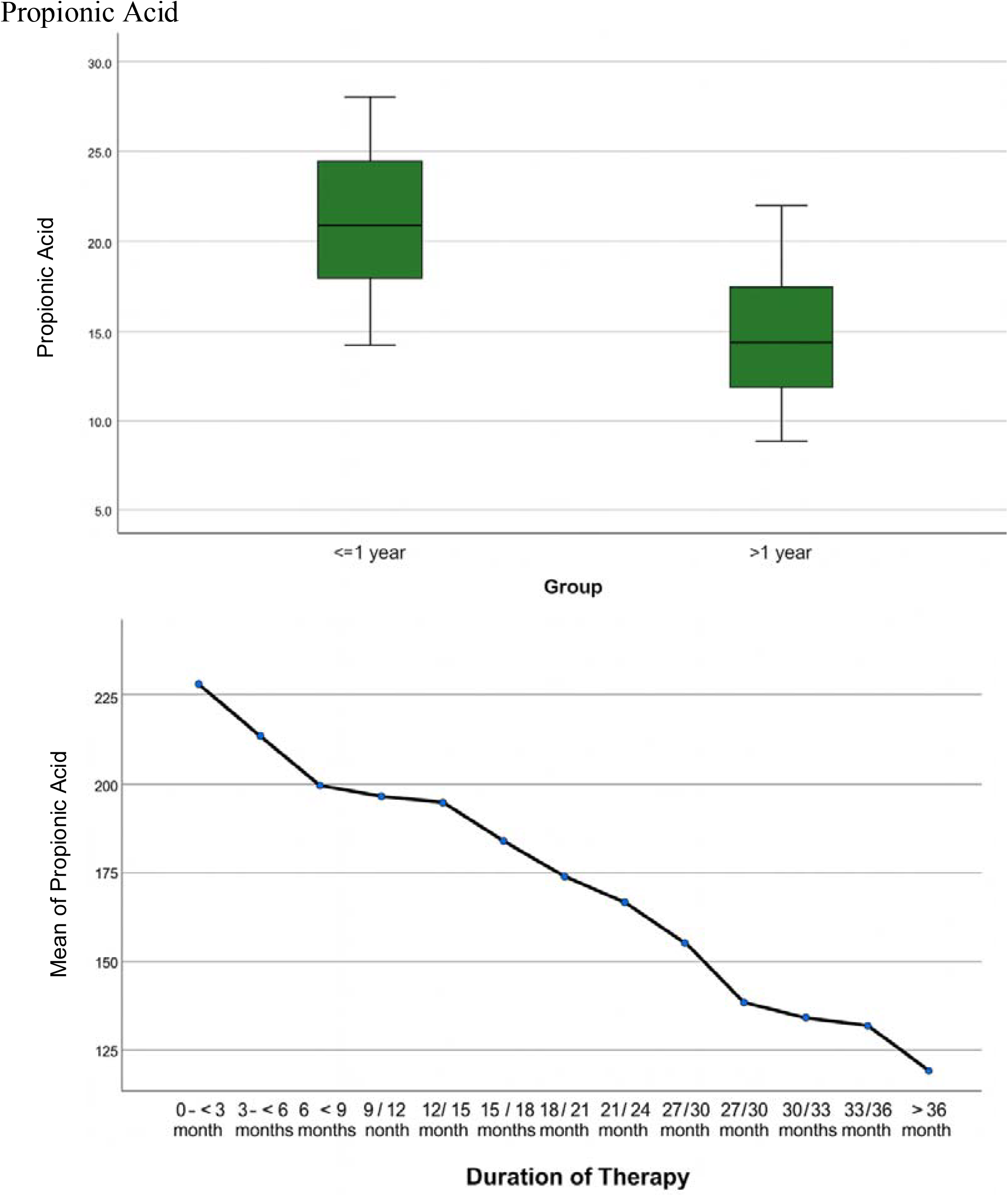
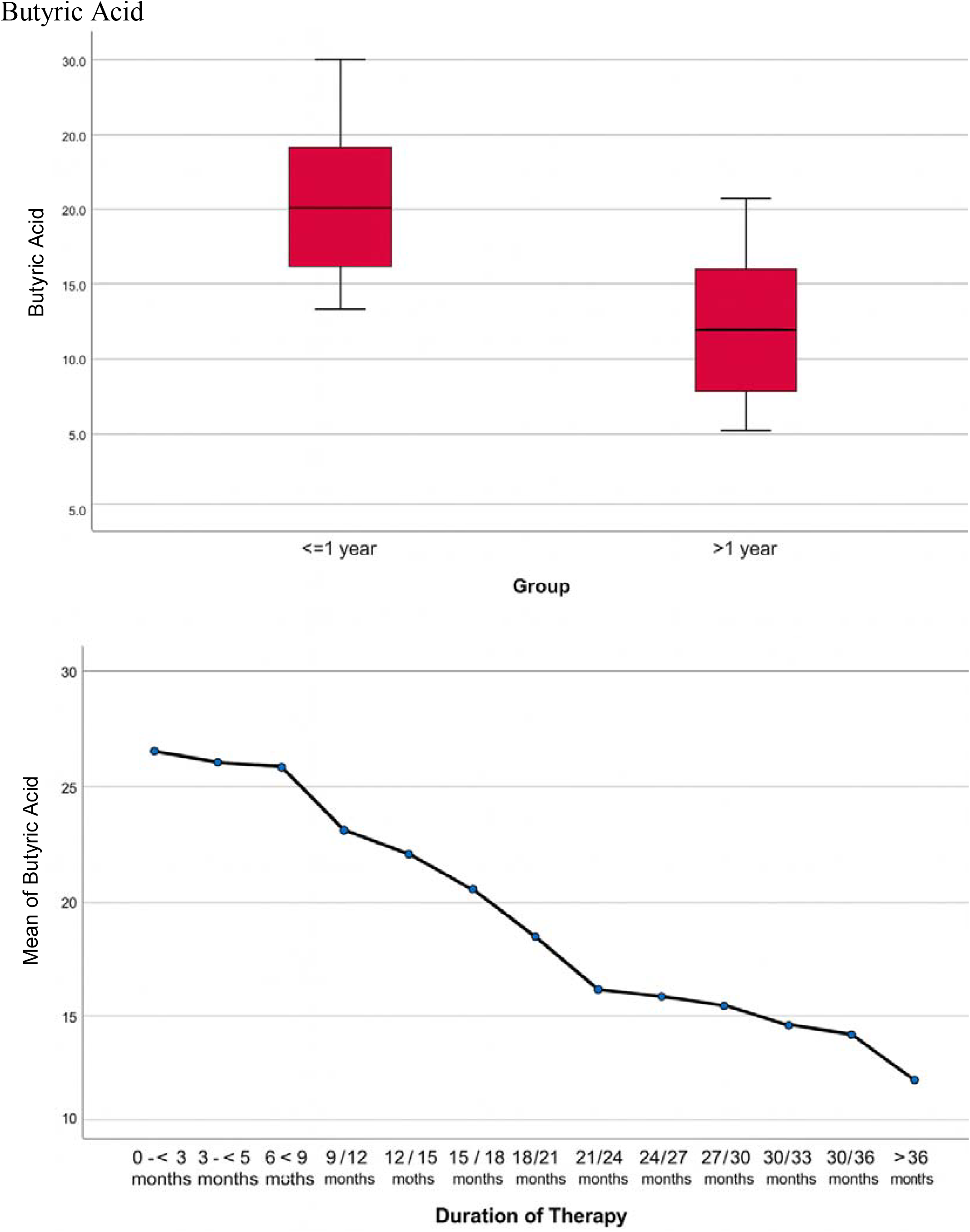
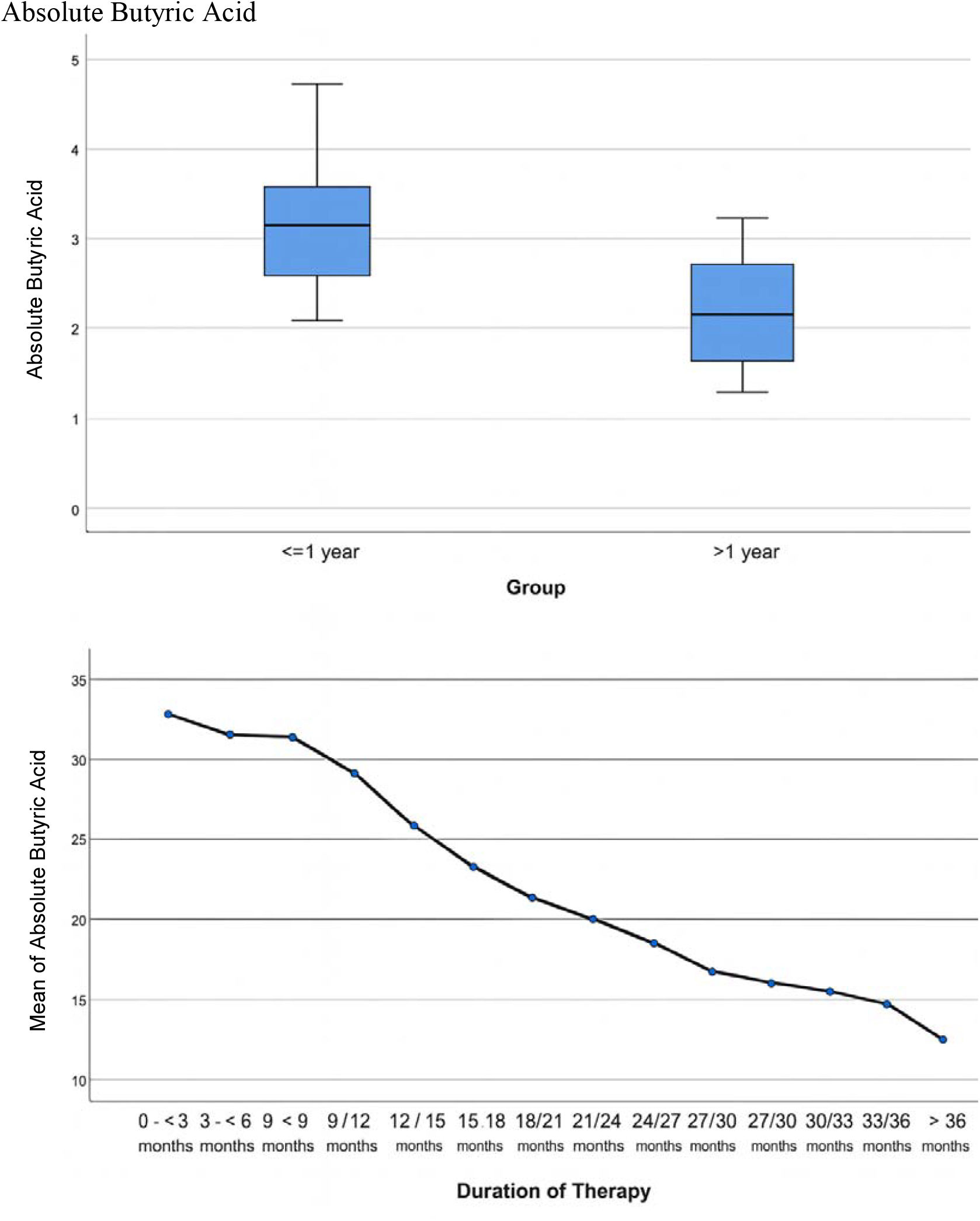
SCFA Profile in Pediatric Epilepsy Patients Based on Duration of Valproic Acid Therapy.

**Figure 2.**
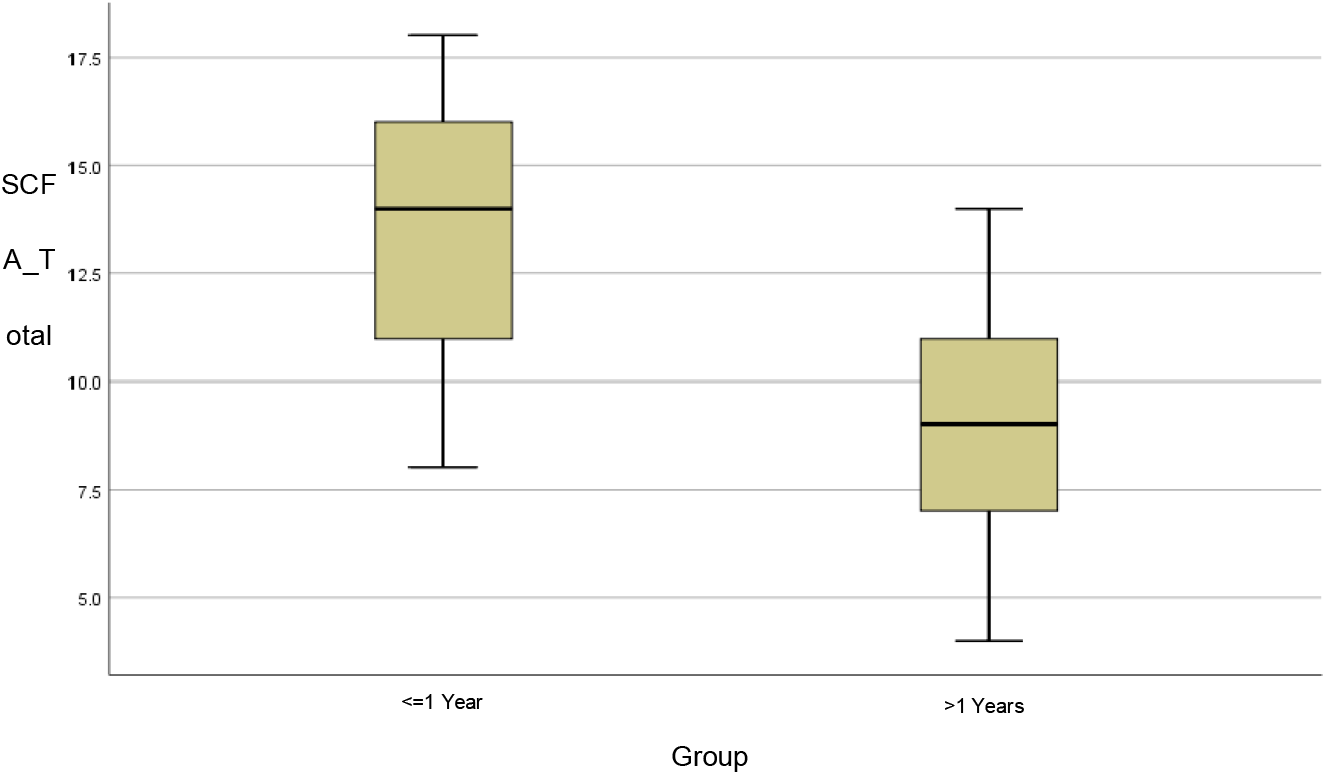
Box Plot of Differences in Total SCFA Levels Based on Duration of Therapy.

**Figure 3.**
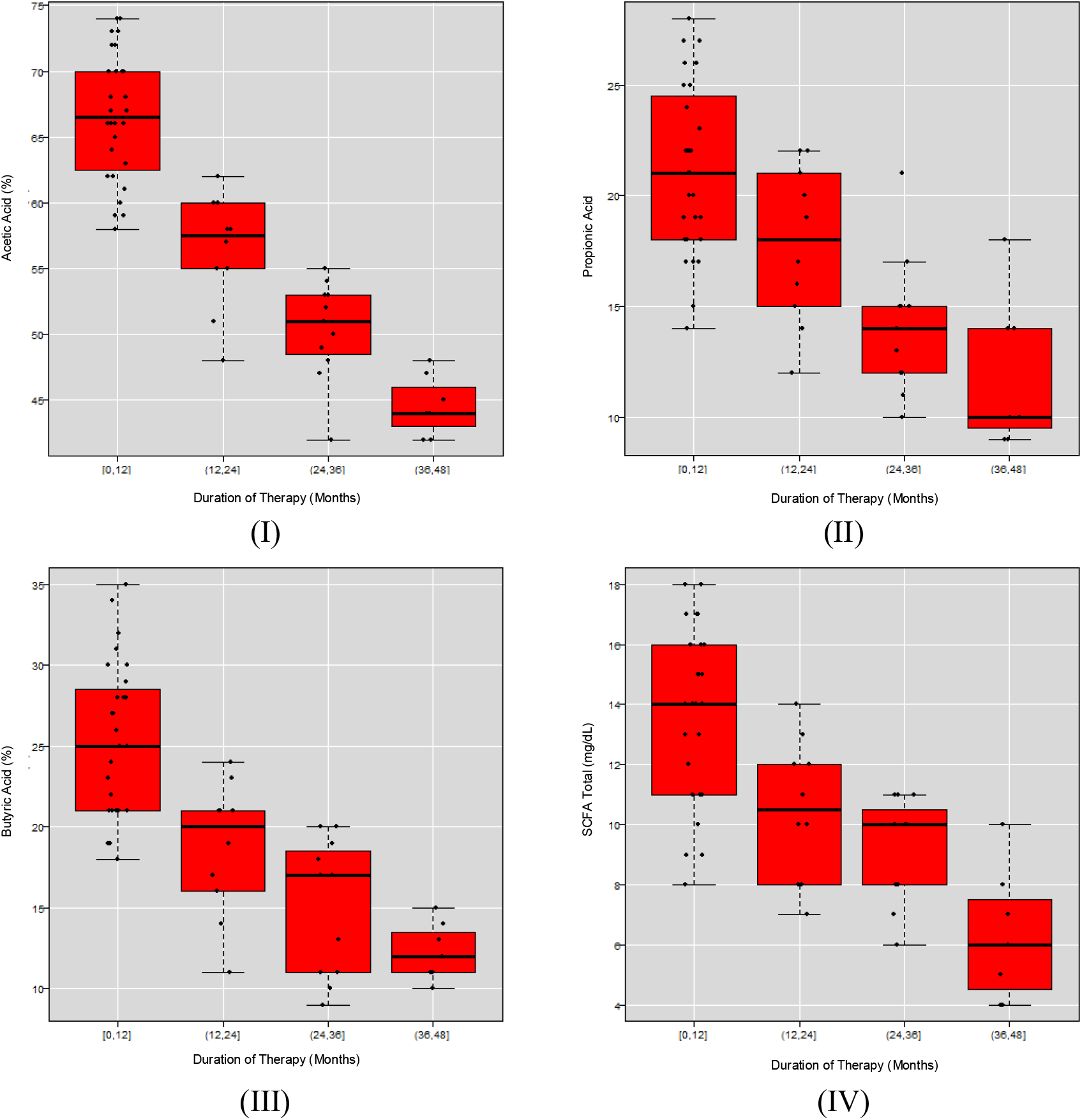
Distribution of Short Chain Fatty Acid Levels According to Duration of Therapy.

Analysis of SCFA within each group showed that among children receiving therapy ≤1 year, concentrations of acetate, propionate, and butyrate showed a declining trend from 0 to 12 months, although not statistically significant. In contrast, in the >1 year group, SCFA concentrations decreased more sharply. Acetate levels decreased significantly from 60.0% at 12–<15 months to 44.57% after 36 months. A similar pattern was observed for propionate, butyrate, and total SCFA.

P-values for acetate (p = 0.015), propionate (p = 0.002), butyrate (p = 0.040), and total SCFA (p = 0.014) indicated statistically significant declines during therapy durations >1 year. Post-hoc Bonferroni analysis confirmed significant pairwise differences in acetate levels between multiple duration groups (e.g., 12–<15 vs. >36 months, p = 0.001).

Between-group comparisons revealed that children treated for 0–<3 months had significantly higher acetate levels than those treated for >36 months (mean difference: 23.72, p<0.001). Similar findings were observed across other duration categories. Propionate and butyrate levels also declined significantly over time, with the largest differences observed in comparisons involving the >36-month group. Absolute butyrate and total SCFA levels followed the same declining trend with statistically significant differences between short and long duration groups.

Multivariate logistic regression analysis indicated that total SCFA was significantly associated with the duration of valproic acid therapy. Children with lower total SCFA had higher odds of being in the >1 year therapy group (OR: 0.49, 95% CI: 0.33–0.73, p<0.001). Other covariates such as age and seizure duration were not significantly associated. Seizure type (generalized vs. focal) showed a significant association (OR: 7.21; p = 0.046).

## DISCUSSION

Short-chain fatty acids (SCFAs) are the primary fermentation products of gut microbiota and play an essential role in human health, including the function of the central nervous system. The main components of SCFAs include acetic acid, propionic acid, and butyric acid, all of which are known to influence various neurophysiological aspects, including in patients with epilepsy [29].

This study employed a cross-sectional approach to evaluate differences in SCFA levels between two groups of pediatric epilepsy patients: those receiving valproic acid therapy for less than one year and those undergoing therapy for more than one year. A total of 56 participants were evenly divided into these two groups (n = 28 in each).

Sex distribution showed a majority of male patients (53.6%), consistent with epidemiological data indicating a slightly higher incidence of epilepsy in males, with a peak during the first year of life and a gradual decline with increasing age, especially after 10 years. This pattern is more prominent in low-to middle-income countries due to limited access to healthcare services and demographic factors [29].

Most patients in this study experienced generalized seizures (71.4%). Although global studies suggest focal seizures are the most common type in both pediatric and adult populations [Beghi E], generalized tonic-clonic seizures remain dominant in regions with lower socioeconomic status [30]. Additionally, the majority of seizures lasted less than 2 minutes (71.4%), which is consistent with the findings of Larsen et al., who reported that most seizures lasted under two minutes, with average durations of 79 seconds for generalized seizures and 103 seconds for focal seizures [31].

The types of SCFAs analyzed in this study included acetate, propionate, and butyrate, which physiologically represent the dominant SCFA components in the gastrointestinal tract. In a healthy gut microbiota environment, SCFA composition typically consists of acetate (40–75%), propionate (9–29%), and butyrate (9–37%), with total SCFA concentrations ranging from 4–18 mg/mL. In this study, mean SCFA levels in all subjects remained within the normal range; however, the group that received valproic acid therapy for more than one year demonstrated lower SCFA concentrations compared to the group treated for less than one year.

Studies on SCFA levels in pediatric epilepsy patients remain limited. Research by Zhong et al. comparing fecal SCFA levels between valproic acid-treated children and controls reported significant reductions in SCFAs, including acetate, propionate, and butyrate. This was accompanied by decreased populations of *Bifidobacterium spp*., *Lactobacillus spp*., and *Limosilactobacillus spp*., along with increased levels of *Ligilactobacillus spp*. and *Prevotellaceae spp*. [32].

Long-term use of valproic acid is known to affect the gut microbiota composition, which plays a critical role in SCFA production. A three-month cohort study on epilepsy patients indicated that valproic acid therapy did not significantly alter overall gut microbiota diversity. Nonetheless, an increased Firmicutes to Bacteroidetes ratio was observed after three months of therapy [24].

### Regression Analysis

Linear regression analysis indicated that each additional month of valproic acid therapy was associated with a decrease in acetic acid concentration by approximately 0.6% (~7% per year), and in propionic and butyric acids by 0.26% and 0.37% per month, respectively (~3–5% per year), after adjusting for dose and body weight. Total SCFA levels declined by an average of 0.18 mg/dL per month (2.16 mg/dL per year), with a 95% confidence interval of 0.13–0.22 mg/dL. This modeling confirmed a linear relationship between treatment duration and SCFA decline, despite the sample not being randomly selected.

This study demonstrates a significant difference in SCFA levels among children with epilepsy based on the duration of valproic acid therapy, suggesting a potential time-dependent impact of the drug on gut microbiota and its metabolic products. The observed decline in SCFA concentrations, particularly acetate, butyrate, and branched-chain SCFAs such as isobutyrate and isovalerate, in the long-term treatment group supports the hypothesis that valproic acid contributes to gut microbiota dysbiosis. These findings are consistent with previous experimental studies in animal models, where valproic acid administration led to alterations in microbial composition and a reduction in fecal SCFA concentrations [32–34].

Zhou et al. reported that untreated pediatric epilepsy patients exhibited increased abundance of phylum *Actinobacteria* and genera *Escherichia/Shigella, Collinsella, Streptococcus*, and *Megamonas* compared to healthy controls [35]. This reflects gut flora alterations that may underlie or exacerbate epileptic conditions. However, following three months of antiepileptic therapy, no significant differences were found between pre- and post-treatment groups, except for a reduction in *Actinobacteria*, suggesting that valproic acid-induced microbiota changes are not rapidly reversible and may require a longer duration to manifest clinically or molecularly relevant effects.

Long-term use of valproic acid has also been associated with gut dysbiosis, as described by Gong et al., who observed an increased presence of opportunistic pathogenic genera and decreased microbial diversity in epilepsy patients [36]. The reduction in SCFA levels with prolonged therapy is a matter of concern, as SCFAs are crucial for maintaining intestinal mucosal integrity, modulating the immune system, and supporting neurological function through the gut-brain axis [37,38]. A sustained decrease in SCFAs may impair antiepileptic drug absorption, increase intestinal permeability, and trigger systemic inflammation, all of which could exacerbate seizure severity.

Physiologically, valproic acid enhances gamma-aminobutyric acid (GABA) levels, the principal inhibitory neurotransmitter in the brain. However, its metabolic side effects, including impaired mitochondrial β-oxidation, increased reactive oxygen species (ROS), and disturbances in fatty acid metabolism, have been extensively documented [39]. Moreover, valproic acid acts as a histone deacetylase (HDAC) inhibitor, which may influence microbial gene expression and metabolite production, including SCFAs. A study by Poolchanuan et al. found that high concentrations of VPA (100 µM) could increase trans-9-elaidic acid production while inhibiting biosynthesis of other fatty acids by gut microbes and model fungi, highlighting VPA’s direct impact on microbial metabolism and intestinal fatty acid profiles [39].

Animal studies have further demonstrated a reduction in neurotransmitters such as tryptophan, kynurenine, and 5-HIAA in VPA-treated groups compared to controls [32]. These decreases correlated with the abundance of genera such as *Pseudomonas, Collinsella*, and *Streptococcus*, as well as lower SCFA concentrations, reinforcing the connection between gut microbiota, SCFA metabolites, and neurotransmitter homeostasis. This interplay underscores the central role of the gut-brain axis in neurological disorders like epilepsy.

Clinically, the decline in SCFAs may impair antiepileptic therapy effectiveness. SCFAs not only support gut health but also influence mucosal immune responses, inflammation regulation, and drug metabolism [37,40]. In a state of dysbiosis, these functions may be compromised, potentially leading to suboptimal therapeutic response and increased seizure frequency in pediatric patients [38,40].

Nevertheless, the findings of this study must be interpreted cautiously due to several limitations. Dietary variability among children, which is difficult to control, can significantly influence SCFA concentrations [37]. Dietary fiber content, food types, and eating patterns directly affect SCFA-producing microbiota. Moreover, comorbid conditions, concurrent medication use, and environmental factors may act as confounding variables. Therefore, future studies with dietary control and longitudinal observation are needed to strengthen these findings.

This study is the first to specifically compare total SCFA levels based on the duration of valproic acid therapy in pediatric epilepsy patients. Unlike previous research that primarily explored general metabolic effects of valproic acid or used animal models, this study seeks to determine the critical time point at which SCFA levels begin to decline significantly. By stratifying therapy duration at the 3-month mark, the study aims to identify a clinically relevant threshold for microbiota alteration.

These findings open potential avenues for new therapeutic strategies that incorporate gut microbiota health as an integral part of long-term epilepsy management. Gut-supportive interventions such as high-fiber diets, probiotics, or prebiotics may serve as beneficial adjuncts to maintain SCFA levels and therapeutic efficacy. In the future, integrating pharmacological and microbiological approaches may enhance the quality of life for children living with epilepsy.

## CONCLUSION

Based on the findings of this study, it can be concluded that the total levels of short-chain fatty acids (SCFAs), including acetic acid, butyric acid, and propionic acid, were higher in pediatric epilepsy patients receiving valproic acid therapy for less than one year compared to those undergoing therapy for more than one year. These results suggest that the duration of valproic acid treatment affects SCFA production, which plays a crucial role in maintaining gut mucosal resilience. The observed differences in SCFA concentrations may serve as potential indicators for assessing intestinal mucosal integrity in children with epilepsy according to the duration of therapy.

## Institutional Review Board Statement

Not applicable.

## Ethical Statement and Informed Consent Statement

This study was approved by the Ethics Committee of Prof.□Dr.□R.□D.□Kandou Manado Central General Hospital (Approval No. 022/EC/KEPK□KANDOU/1/2024), and written informed consent was obtained from all participants.

## Data Availability Statement

The datasets used and/or analysed during the current study available from the corresponding author on reasonable request.

## Acknowledgments

Not applicable.

